# Risk Assessment Techniques and Risk Management Practices in Healthcare: A Comparative Survey of the United States and United Kingdom

**DOI:** 10.64898/2026.03.10.26348078

**Authors:** Eugenia O’Kelly, James Ward, P. John Clarkson

**Affiliations:** Cambridge University; University of Cambridge

**Keywords:** Healthcare risk management, Patient safety, Risk assessment, Root Cause Analysis, Failure Modes and Effects Analysis, United States, United Kingdom

## Abstract

**Introduction and Aims:** Effective risk management is essential for improving patient safety, yet limited empirical evidence exists on how risk assessment techniques are used in routine healthcare practice. This study examines current risk management practices in healthcare organisations and compares approaches used in the United States (USA) and the United Kingdom (UK).

**Methods:** A survey of practising healthcare risk managers in both countries examined the use of risk assessment techniques and organisational risk management practices, including team composition and perceived organisational resources. A total of 160 risk managers from the USA and 40 from the UK, representing a wide range of clinical and administrative healthcare areas, participated in the study.

**Results:** Root Cause Analysis (RCA) was the most frequently used risk assessment technique in the USA, followed by Failure Modes and Effects Analysis (FMEA). In the UK, risk matrices were most commonly used, followed by Strengths, Weaknesses, Opportunities, and Threats (SWOT) analysis. Risk managers in both countries preferred prospective risk management; however, organisational practice in the USA was reported to be significantly more retrospective. Approximately half of respondents reported insufficient organisational resources for effective risk management, most commonly limited time and staffing. In addition, only 43% of USA respondents and 47% of UK respondents reported that recommended risk improvement measures were implemented more than half of the time.

**Conclusions:** Healthcare risk management practices differ between the USA and the UK, particularly in the techniques used, organisational support, and the balance between prospective and retrospective approaches. Strengthening institutional support, increasing multidisciplinary participation, and expanding the use of structured prospective techniques may improve the effectiveness and maturity of healthcare risk management.

**Key Message (BMJ Requirement):** *What is already known on this topic?:* Empirical studies examining how risk management is conducted in healthcare organisations are limited. Most knowledge about healthcare risk management practice in the United States and the United Kingdom is inferred from guidelines or recommended best practices rather than from studies of actual practice. Evidence regarding risk management practice in UK healthcare institutions is particularly scarce.

*What this study adds:* This study provides empirical evidence on the current state of healthcare risk management practice in the USA and the UK. It identifies the risk assessment techniques most commonly used in healthcare organisations and examines how risk management activities are conducted. The findings highlight strengths and limitations in current practice and provide a comparative perspective on risk management approaches across the two healthcare systems.

*How this study might affect research, practice, or policy:* The study identifies several areas where healthcare risk management practice may be strengthened, including organisational support for risk management, multidisciplinary participation in risk assessment, and implementation of risk improvement measures. The findings also provide insight into the adoption of risk assessment techniques within healthcare, informing future research, policy development, and efforts to improve patient safety management practices.

## Introduction

The importance of effective healthcare risk management in enabling safe and efficient medical practice has been widely recognized [1–3]. However, comparatively little empirical research has examined the techniques and practices currently used by hospitals to manage risk. This paper addresses two key areas: (1) the risk assessment techniques most commonly used in healthcare risk management, and (2) current organisational practices associated with their use, including team size and composition, prospective versus retrospective focus, and allocation of resources.

A substantial body of literature proposes or demonstrates the potential application of various risk management techniques in healthcare settings. This includes studies examining established techniques such as Root Cause Analysis (RCA) and Failure Modes and Effects Analysis (FMEA) [4–8], as well as research proposing or developing new techniques for analysing risk [9,10]. Despite this extensive literature, there is limited empirical evidence regarding how frequently these risk assessment techniques are used in routine healthcare practice. For example, some authors describe FMEA as a commonly used technique in healthcare [11,12], whereas others suggest that its use remains limited [13].

Several studies have attempted to identify which risk management techniques are used in healthcare through bibliometric analyses of published academic literature [14–17]. However, bibliometric approaches have limitations when used to infer actual practice. Many studies appearing in the academic literature are conducted by researchers rather than healthcare risk professionals and may reflect the application of novel techniques for research purposes rather than techniques routinely used within healthcare organisations. As a result, bibliometric analyses may overrepresent new or experimental approaches that are of interest to researchers.

Conversely, techniques widely used by healthcare managers may receive limited attention in the academic literature if they are considered routine or lack novelty.

Other studies have inferred the use of particular techniques based on their recommendation by authoritative organisations, such as the Institute for Safe Medication Practices [18], or based on broader support from policy makers [19]. While these approaches can help identify widely recognised techniques, they do not provide direct empirical evidence regarding their actual use in healthcare organisations.

This study provides empirical data on the use of risk assessment techniques by practising healthcare risk managers. In addition, the study examines how these techniques are implemented within organisations, including aspects of organisational structure, team composition, and the balance between prospective and retrospective risk management activities.

Comparative research examining risk management maturity across industries has suggested that healthcare risk management may be less mature than risk management practices in other safety-critical sectors such as aerospace, oil, and gas [20–22]. By examining current risk management practices in hospitals in the United States and the United Kingdom, this study aims to provide insight into the current state of practice and potential factors that may contribute to differences in maturity.

Conducting empirical research on healthcare risk management practice presents methodological challenges. Previous studies have noted the difficulty of identifying healthcare risk managers in the United Kingdom and have reported low response rates when surveying this population [23,24]. Individuals responsible for risk management within the National Health Service (NHS) are often difficult to identify externally because job titles vary substantially between hospitals and frequently do not explicitly reference risk management responsibilities. Risk management functions may be carried out by individuals with diverse administrative or clinical backgrounds depending on the structure of the hospital. No comprehensive hospital-level or NHS-wide database of risk management personnel was identified for this study, and a freedom of information request did not provide even an approximate estimate of the number of individuals performing risk management roles.

## Methods

An original survey instrument was developed to collect data on healthcare risk management practices. The instrument consisted of three categories of questions: (1) demographic questions,(2) questions regarding the use of risk assessment techniques, and (3) questions relating to organisational practices in risk management.

A key step in developing the survey was constructing a list of risk assessment techniques likely to be used in healthcare organisations. The initial list of techniques was derived from BS IEC 31010:2019 and a review of scholarly and grey literature. The list was subsequently refined through consultation with prior research conducted by the Healthcare Design Group [25–29].

A final list of twelve risk assessment techniques and technique categories was included in the survey instrument, with an option for respondents to enter and rank additional techniques not listed. The list included both established techniques commonly mandated or recommended by oversight bodies as well as newer system-oriented techniques proposed for healthcare safety analysis. During survey development, feedback was obtained from the Head of Risk and Quality at a large teaching hospital and from a pilot group of ten healthcare risk managers. The final list of techniques included in the survey was:

- Root Cause Analysis (RCA)
- Failure Modes and Effects Analysis (FMEA), Healthcare Failure Modes and Effects Analysis (HFMEA), and/or Failure Modes, Effects, and Criticality Analysis (FMECA)
- Event Tree Analysis (ETA), Fault Tree Analysis (FTA), or Bow-tie analysis (combined ETA and FTA)
- Risk matrix
- Significant Event Audit (SEA)
- Critical Incident Technique (CIT)
- Hazard and Operability Analysis (HAZOP)
- Human Reliability Assessment (HRA) techniques, including Human Error Assessment and Reduction Technique (HEART), Paired Comparison (PC), Technique for Human Error Rate Prediction (THERP), and the Systematic Human Error Reduction and Prediction Approach (SHERPA)
- Strengths, Weaknesses, Opportunities, and Threats (SWOT) analysis
- Structured What-If Technique (SWIFT)
- Functional Resonance Analysis Method (FRAM)
- System-Theoretic Process Analysis (STPA)

Feedback from pilot participants resulted in the inclusion of the risk matrix and SWOT analysis. Although risk matrices are primarily visualisation tools, previous research indicates that they are frequently used within healthcare risk management practice [23,30]. Similarly, SWOT analysis is not specifically designed as a risk assessment technique but has been recommended or applied for this purpose in some risk management contexts [30–34].

Two versions of the survey instrument were produced: one for respondents in the United Kingdom and one for respondents in the United States. Both versions contained identical questions but used country-specific terminology and spelling where appropriate (e.g., A&E in the UK and ER in the USA).

Ethical approval for the study was obtained from the University of Cambridge Department of Engineering Research Ethics Committee and a local NHS trust (PRN 8443).

Participation in the study was restricted to individuals performing healthcare risk management activities. For the purposes of this research, a risk manager was defined as an individual who regularly conducts risk management activities, defined as “the process of investigating, analysing, or making judgements about an incident or potential risk”. Guidance was provided to survey recipients to allow them to self-assess their eligibility. Screening questions were used to exclude clinical staff and administrators who did not routinely conduct risk management activities. Eligible activities included:

- Investigating incidents
- Formally identifying risks
- Assessing or evaluating risks (quantitatively or qualitatively)
- Planning measures to control or treat risks
- Reviewing risk controls
- Monitoring risks and threats

Clinical staff whose involvement was limited to submitting incident reports or evaluating clinical risk on a patient-by-patient basis were considered ineligible for participation.

In the United Kingdom, survey invitations were distributed through professional groups where NHS risk managers are likely to participate, including NHS quality management forums and the Institute of Risk Management’s “Health and Care” special interest group. UK responses were collected between 12 September 2019 and 17 November 2020. A total of 54 individuals completed the survey, of whom 40 met the eligibility criteria.

Healthcare risk management has a more established professional profile in the United States than in the United Kingdom. In the USA, risk management can be studied through specialised degree programmes and supported through professional certification. The American Society for Health Care Risk Management (ASHRM), established in 1980, represents one of the largest professional organisations for healthcare risk managers and currently includes approximately 6,000 members. ASHRM also administers the Certified Professional in Health Care Risk Management (CPHRM) certification programme.

To recruit respondents in the United States, an invitation to participate in the survey was distributed through the ASHRM mailing list, which included approximately 2,000 individuals. Data from US respondents were collected between 12 September 2019 and 21 November 2019. A total of 191 responses were received, of which 160 met the eligibility criteria.

## Results

Participants in the study were involved in a wide range of hospital risk management activities. Most respondents (85% of USA respondents and 67.5% of UK respondents) reported conducting risk management activities across all or most clinical areas. A smaller proportion (22.5% of UK respondents and 12.5% of USA respondents) reported working in one or more specific clinical areas. In addition, 80% of UK respondents and 45% of USA respondents indicated that they were responsible for risk management related to administrative, operational, employee management, or other non-clinical hospital activities.

### Familiarity with Risk Assessment Techniques

Participants were asked to identify risk assessment techniques with which they were familiar, regardless of whether they had personally used them. Figure 1 shows participant familiarity with the twelve technique categories included in the survey.

**Figure 1:**
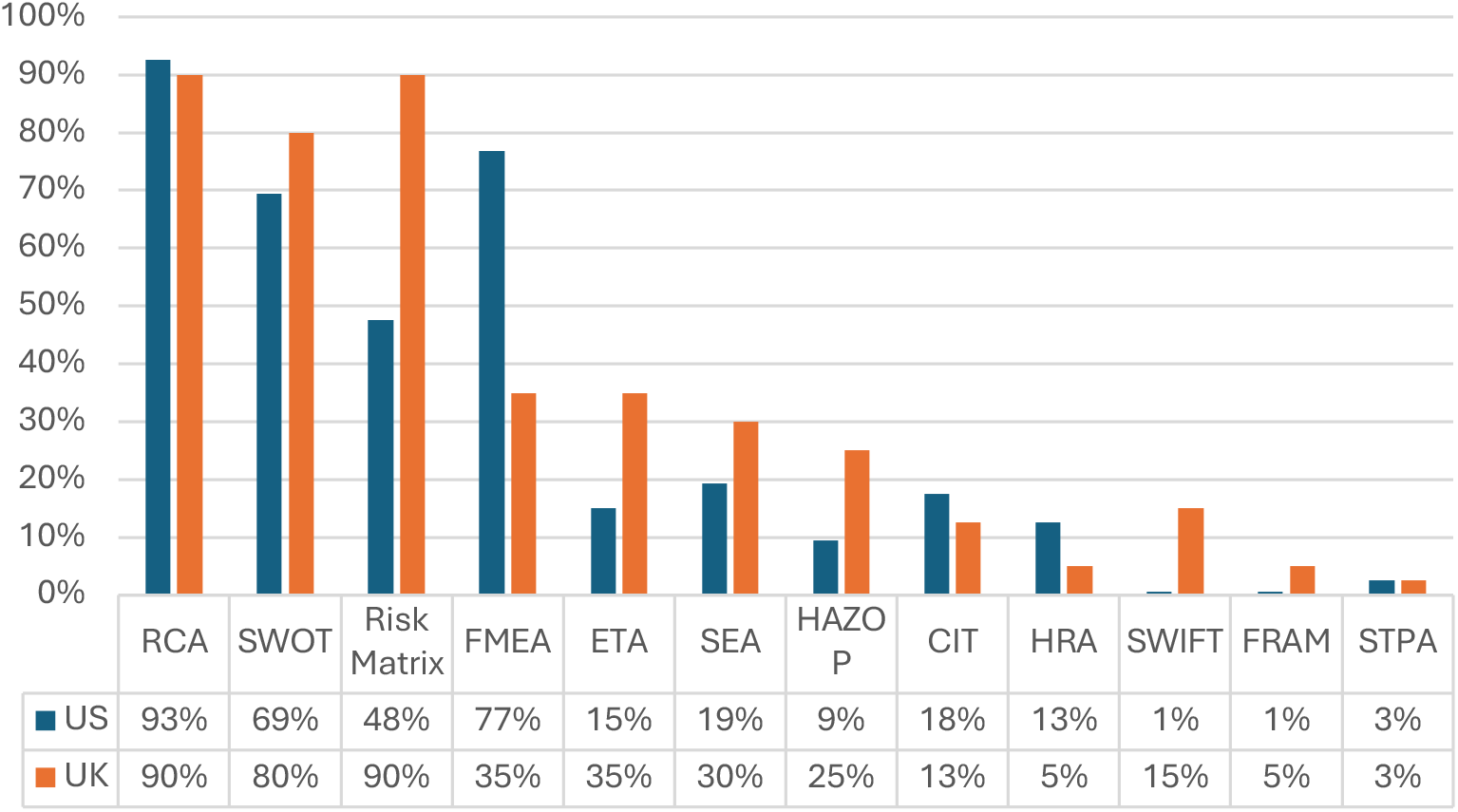
Percentage of participants familiar with each risk assessment technique.

Two techniques showed high familiarity among both USA and UK respondents: Root Cause Analysis (RCA) and SWOT analysis. RCA was the most widely recognised technique among respondents, with 93% of USA risk managers and 90% of UK risk managers reporting familiarity. The second most familiar technique among USA respondents was Failure Modes and Effects Analysis (FMEA), with 77% reporting familiarity. In contrast, only 35% of UK respondents reported familiarity with FMEA.

Both USA and UK respondents also reported familiarity with SWOT analysis, with 69% of USA respondents and 80% of UK respondents indicating familiarity. UK respondents reported particularly high familiarity with the risk matrix technique, with 90% indicating familiarity.

Several techniques designed to analyse socio-technical systems or human reliability were less familiar to respondents. Significant Event Audit (SEA) was recognised by 19% of USA respondents and 30% of UK respondents. Human Reliability Assessment (HRA) techniques were recognised by 13% of USA respondents and 5% of UK respondents. Newer risk assessment techniques, such as System-Theoretic Process Analysis (STPA) and the Functional Resonance Analysis Method (FRAM), were rarely recognised by respondents.

### Frequency of Risk Assessment Technique Use

Participants were asked to indicate how frequently they used each technique in their professional practice using a five-point Likert scale: “never”, “infrequently”, “about half the time”, “most of the time”, and “always”. For visual representation, responses were converted to a numerical scale ranging from 0 (“never”) to 4 (“always”). Figure 2 illustrates the relative frequency of technique use among USA and UK respondents.

**Figure 2:**
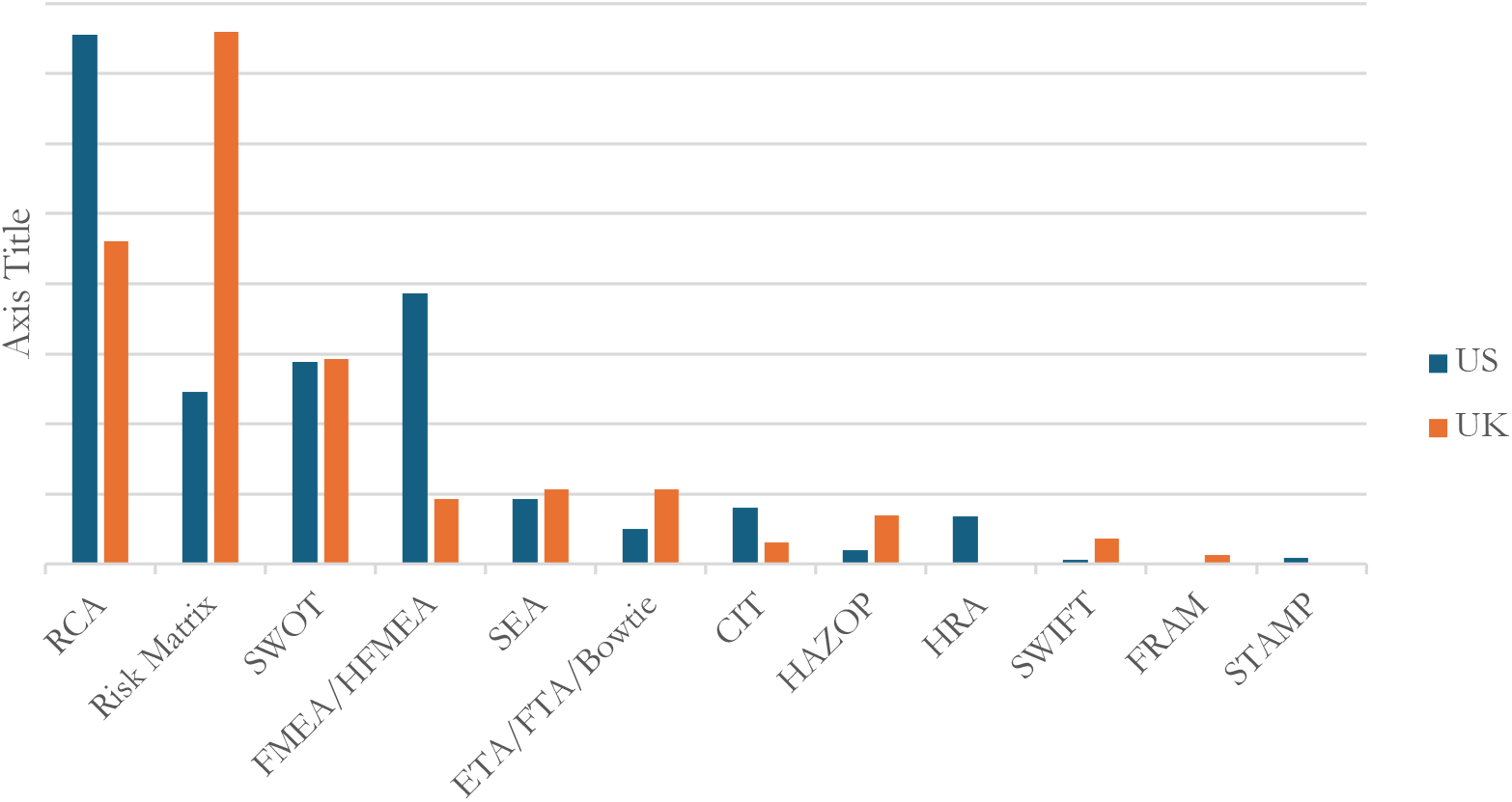
Relative frequency of risk assessment technique use based on weighted Likert-scale responses.

RCA was the most frequently used technique among USA respondents, with 57% reporting that they used RCA at least half of the time. In contrast, RCA was used less frequently among UK respondents: only 3% reported using the technique “most of the time” or “always”, while 55% reported that they “never” used RCA.

The second most frequently used technique among USA respondents was FMEA or HFMEA. However, only 17% reported using the technique “always” or “most of the time”, and 13% reported using it “about half the time”. The majority of respondents (70%) reported that they used FMEA/HFMEA “never” or “infrequently”. Use of FMEA/HFMEA was less frequent among UK respondents, with 95% reporting that they “never” or “infrequently” used the technique.

Among UK respondents, the most frequently used technique was the risk matrix, with 65% reporting that they used it “always” or “most of the time”. SWOT analysis was the next most frequently used technique, with 23% reporting use “always” or “most of the time”. These techniques were less frequently used among USA respondents, with 13% reporting frequent use of the risk matrix and 8% reporting frequent use of SWOT analysis.

Some techniques were reported as being used at least half of the time by more than 10% of respondents in at least one country. For example, Event Tree Analysis (ETA) was reported as being used at least half of the time by 4% of USA respondents and 13% of UK respondents. However, only a small proportion of respondents reported using ETA “most of the time” or “always”.

Several techniques were rarely used by respondents. All USA and UK respondents reported that they never used FRAM. STPA was reported as being used by 1% of USA respondents and 2% of UK respondents. Among the human reliability techniques, the Human Error Assessment and

Reduction Technique (HEART) was the most frequently reported, although 94% of USA respondents and 98% of UK respondents indicated that they used it “never” or “infrequently”.

### Retrospective and Prospective Risk Assessment Preferences

Respondents in both countries indicated a preference for proactive approaches to risk management. Among USA respondents, 76.4% indicated that healthcare risk management should be mostly or entirely proactive. Similarly, 70% of UK respondents indicated a preference for mostly or entirely proactive risk management.

However, respondents reported that current organisational practice was more retrospective in nature. Among USA respondents, 65% reported that risk management in their organisation was “mostly reactive” or “entirely reactive”. In contrast, 27% of UK respondents reported that risk management in their organisation was mostly or entirely reactive. Conversely, only 7% of USA respondents reported that risk management was mostly or entirely prospective, compared with 27% of UK respondents.

A chi-squared test indicated a statistically significant association between country and the reported orientation of risk management practice, with UK respondents reporting more prospective approaches and USA respondents reporting more retrospective approaches (p < 0.001).

### Resources for Risk Management

Participants were asked whether their organisation provided sufficient resources for risk management activities. Among USA respondents, 54% reported that sufficient resources were available, compared with 43% of UK respondents. Figure 3 illustrates the specific resources that respondents reported as lacking.

**Figure 3:**
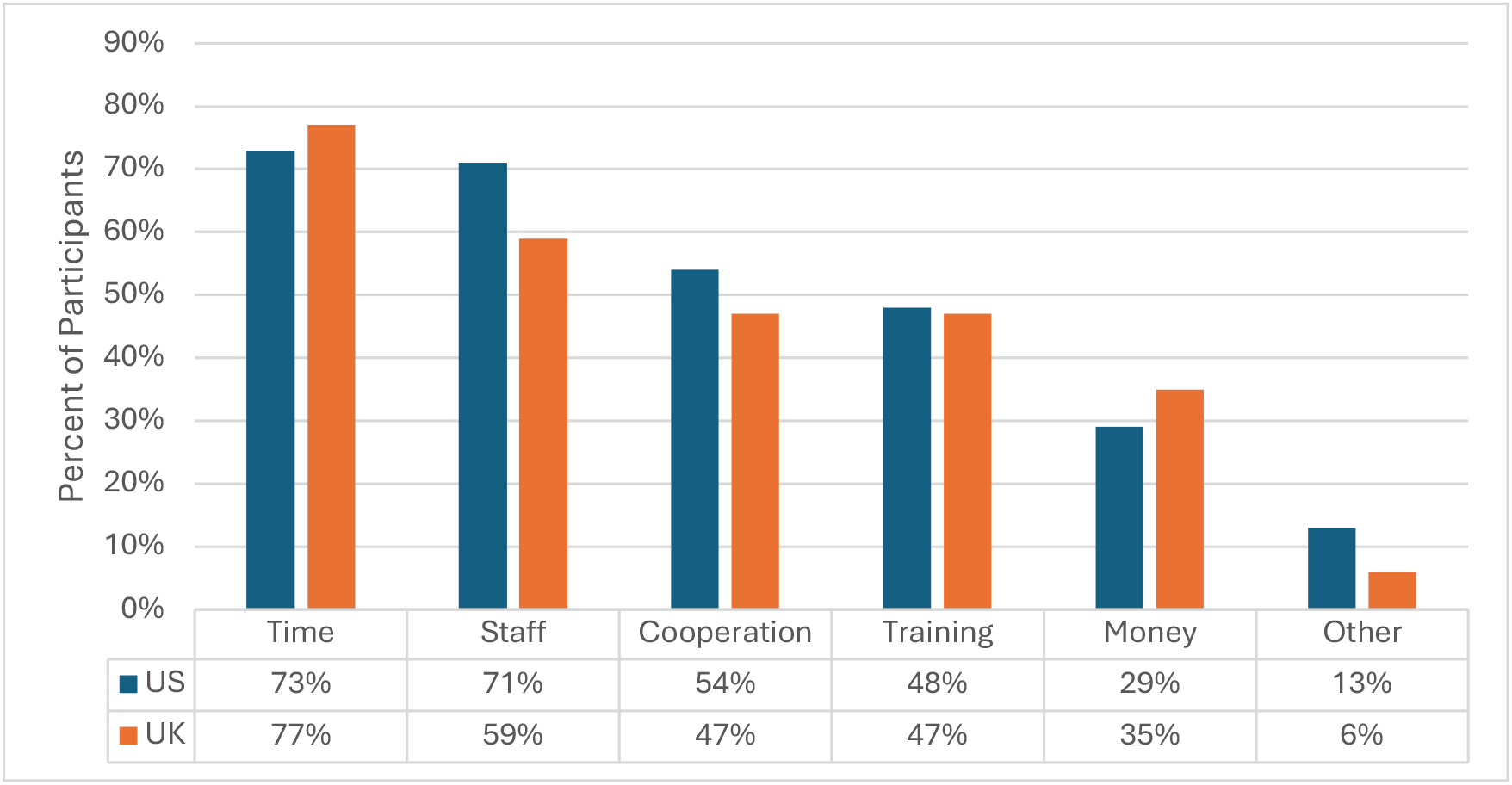
Resources reported as lacking by respondents who indicated insufficient organisational support.

**Figure 4:**
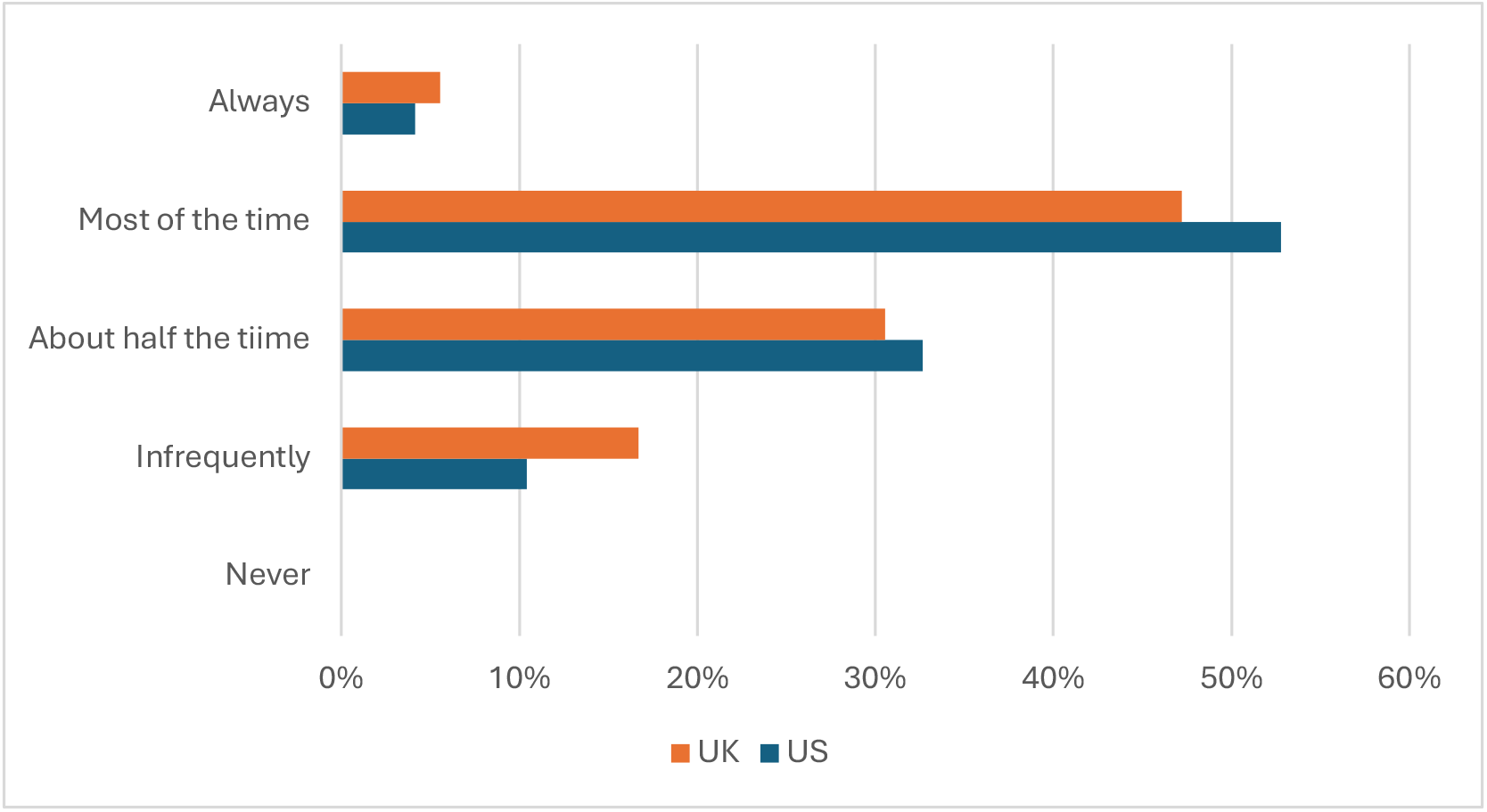
How often risk treatment recommendations were implemented according to US and UK risk managers

Respondents in both countries identified similar resource constraints. The resource most frequently reported as lacking was time, followed by insufficient staffing and limited cooperation from clinical and non-clinical staff.

Guidance from professional standards and organisations recommends that risk assessments be conducted by multidisciplinary teams including representatives from relevant parts of the system [16,35–39]. Respondents were therefore asked whether risk analysis in their organisation was conducted by teams.

Among USA respondents, 87.9% reported that risk analysis was conducted by a team, compared with 82.5% of UK respondents. Team size differed between the two countries. Among USA respondents, 34.4% reported that risk assessment teams typically included five or more individuals. The next most common team size was four members (28.2%), followed by three members (22.9%), meaning that 85.5% of USA teams consisted of three or more individuals.

Among UK respondents, teams were generally smaller, with 66.6% reporting that teams consisted of two or three individuals.

Respondents were also asked whether risk assessment teams included appropriate clinical representatives. In both countries, respondents reported that clinical representatives were included “most of the time” (55.7% of USA respondents and 50% of UK respondents). In contrast, patient representatives were rarely included in risk assessment teams: 92.4% of USA respondents and 86.7% of UK respondents reported that patient representatives were involved “infrequently” or “never”.

Several respondents identified organisational safety culture as a factor influencing risk management practice. Statistical analyses were conducted to examine potential relationships between indicators of organisational safety culture and risk management practices. A chi-squared test using USA data indicated that organisations reporting adequate resources were more likely to implement risk improvement measures (p < 0.01). However, analyses examining relationships between team size and (1) implementation of risk management recommendations or (2) organisational resource sufficiency were not statistically significant (p > 0.05). Similarly, no statistically significant relationship was identified between human factors or ergonomics training and organisational support, team size, inclusion of patient representatives, or inclusion of relevant clinical experts in risk assessment teams.

### Implementation of Risk Management Recommendations

Risk assessments are intended to inform actions that reduce risk within healthcare systems. Respondents were therefore asked how frequently recommendations resulting from risk management activities were implemented within their organisations.

Only 4.2% of USA respondents and 5.6% of UK respondents reported that their recommendations were “always” implemented. A majority reported that recommendations were implemented “most of the time” (52.8% of USA respondents and 47.2% of UK respondents).

However, approximately 43% of USA respondents and 47% of UK respondents indicated that recommended changes were implemented only half of the time or less.

## Discussion

This study identified several differences between risk management practices in the United States and the United Kingdom and highlighted several areas where current practice may be improved. UK respondents reported frequent use of relatively simple risk assessment techniques, particularly the risk matrix and SWOT analysis. While these techniques can support structured discussion of risks, prior research has questioned their reliability when used as the primary method of risk assessment (Ball & Watt, 2013).The frequent use of risk matrices is consistent with previous studies of healthcare risk management practice [25,27]. UK respondents also reported lower levels of organisational support for risk management activities. In addition, risk assessment teams were relatively small, typically consisting of only two or three individuals. Although UK respondents reported somewhat higher levels of participation from clinical staff and patient representatives than their USA counterparts, overall participation remained limited.

Despite these constraints, UK respondents reported that risk management within their organisations was more frequently prospective in orientation, with over 40% indicating that risk management activities were mostly or entirely proactive. However, this finding appears to contrast with the limited reported use of formal prospective risk assessment techniques such as Failure Modes and Effects Analysis (FMEA). One possible explanation is that proactive risk management activities may occur in less formal ways that do not rely on structured techniques. For example, risks may be discussed in operational meetings, safety briefings, or internal reviews without the use of formal analytical frameworks. While such activities may support proactive awareness of risk, the absence of structured techniques may limit the depth, consistency, and reproducibility of the analysis.

In contrast, the results indicate a strong tendency toward retrospective risk assessment in the United States. Although USA respondents expressed a personal preference for proactive risk management approaches, organisational practice was reported to be predominantly reactive. The frequent use of Root Cause Analysis (RCA), which is designed primarily for retrospective investigation of incidents, reflects this orientation.

The relatively infrequent use of FMEA in the USA is notable given national requirements for prospective risk assessment. The National Center for Patient Safety requires prospective risk assessments to be conducted at least annually, and The Joint Commission requires healthcare organisations to prospectively assess at least one high-risk process every eighteen months [39]. The limited reported use of FMEA suggests that prospective risk assessments may be conducted only to meet minimum regulatory requirements. In such cases, prospective risk assessment may function primarily as a compliance activity rather than as a routine component of organisational risk management.

Team structures also differed between the two countries. USA respondents reported larger teams conducting risk assessments, whereas UK teams were typically smaller. Larger teams may provide greater access to diverse expertise and perspectives during risk analysis. However, participation of clinicians and patient representatives remained limited in both countries. In particular, patient representatives were rarely included in risk assessment activities. This is notable given the increasing emphasis on patient engagement in healthcare safety initiatives.

A further finding of this study concerns the implementation of risk management recommendations. The purpose of a risk assessment is to inform actions that reduce or mitigate risk within a system. If the recommendations generated through risk assessment are not implemented, the practical impact of the assessment is likely to be limited [41]. In the present study, only slightly more than half of respondents in both countries reported that recommendations arising from risk assessments were implemented “most of the time.” A substantial proportion of respondents reported that recommendations were implemented only half of the time or less. In the UK sample, 17% of respondents indicated that their recommendations were rarely or never implemented. These findings suggest that barriers to implementation may represent a significant challenge for healthcare risk management practice.

More broadly, the findings highlight a gap between the range of risk assessment techniques proposed in the patient safety literature and those used in routine practice. Techniques designed to analyse socio-technical systems or human reliability, including Human Reliability Assessment (HRA) techniques, the Functional Resonance Analysis Method (FRAM), and System-Theoretic

Process Analysis (STPA), were rarely recognised or used by respondents in either country. This suggests that many techniques discussed in the safety science literature have not yet been widely adopted in healthcare risk management practice.

The study also identified potential relationships between organisational support and the effectiveness of risk management activities. In the USA dataset, organisations reporting sufficient resources for risk management were more likely to implement recommended risk improvement measures. Although no statistically significant relationship was observed between team size and implementation outcomes, organisational resources appear to play an important role in enabling risk management activities to translate into operational improvements.

Several limitations of the study should be considered when interpreting these findings. The sampling method may have influenced the composition of respondents. Survey invitations were distributed through professional groups and organisations associated with healthcare quality and risk management. As a result, the respondents may represent individuals who are more actively engaged in professional networks and continuing education than the broader population of healthcare risk managers.

Many of the organisations through which the survey was distributed provide educational opportunities related to healthcare safety and risk management. These opportunities may include lectures, articles, and discussions introducing new risk assessment techniques or safety concepts. Consequently, the surveyed population may have had greater exposure to emerging risk management approaches than risk managers who are not active in such professional networks.

This potential sampling bias has particular implications for interpreting results related to awareness and use of risk assessment techniques. Respondents who are more actively engaged in professional communities may be more likely to be aware of specialised or recently developed techniques. Conversely, they may also be less likely to report limitations in their own organisational risk management practices. For these reasons, the findings presented in this study may represent a more engaged subset of healthcare risk managers.

Despite these limitations, the study provides empirical insight into the techniques and practices currently used in healthcare risk management. By examining how risk assessment techniques are applied in practice, the findings contribute to a better understanding of the current maturity of healthcare risk management and highlight areas where further development may be required.

## Conclusion

This study examined healthcare risk management practices in the United States and the United Kingdom through a survey of practising risk managers. A total of 160 USA-based and 40 UK-based risk managers representing a wide range of clinical fields reported on their experience with risk assessment techniques and organisational risk management practices.

The findings reveal notable differences between the two countries in the techniques used, the balance between prospective and retrospective risk assessment, team composition, and the implementation of risk management recommendations. In both countries, relatively simple or retrospective techniques were most commonly used, while more structured prospective and socio-technical risk assessment techniques were rarely applied in routine practice.

The results also highlight several areas where healthcare risk management may be strengthened. Respondents in both countries reported limitations in organisational resources, inconsistent implementation of risk management recommendations, and limited participation of clinicians and patient representatives in risk assessment activities. Addressing these challenges may improve the effectiveness of risk management and support the development of more mature safety practices within healthcare organisations.

More broadly, the findings suggest that a gap remains between the range of risk assessment techniques proposed in the safety science literature and those used in routine healthcare practice. Efforts to improve organisational support, strengthen multidisciplinary participation, and integrate structured prospective risk assessment techniques into routine safety management may enhance the ability of healthcare organisations to identify and mitigate risks before patient harm occurs.

## Data Availability

All data produced in the present study are available upon reasonable request to the authors

## Citation

[1] Kohn LT, Corrigan JM, Donaldson MS. To Err is Human: Building a Safer Health System. 2000. 10.17226/9728.

[2] Lammers RL, Willoughby-Byrwa M, Fales WD. Errors and error-producing conditions during a simulated, prehospital, pediatric cardiopulmonary arrest. Simulation in Healthcare 2014;9:174–83. 10.1097/SIH.0000000000000013.

[3] Stelfox HT, Palmisani S, Scurlock C, Orav EJ, Bates DW. The “To Err is Human” report and the patient safety literature. Qual Saf Health Care 2006;15:174–8. 10.1136/qshc.2006.017947.

[4] Almomani MA, Aladeemy M. Lean based approach for identifying risks and improving safety of healthcare delivery systems: A novel application of FMEA and FTA in blood transfusion unit at a local public hospital in Jordan. Proceedings of the International Conference on Industrial Engineering and Operations Management 2017:1103–12.

[5] Faiella G, Parand A, Franklin BD, Chana P, Cesarelli M, Stanton NA, et al. Expanding healthcare failure mode and effect analysis: A composite proactive risk analysis approach. Reliab Eng Syst Saf 2018;169:117–26. 10.1016/j.ress.2017.08.003.

[6] Mohammed AS, Alammari NK, Alabdouli AA, Almansoori DM. On the application of hazard and operability method in patient safety context: opportunities and challenges. Proceedings of the International Conference on Industrial Engineering and Operations Management 2020:1112–9.

[7] Abecassis ZA, McElroy LM, Patel RM, Khorzad R, Carroll C, Mehrotra S. Applying fault tree analysis to the prevention of wrong-site surgery. Journal of Surgical Research 2015;193:88–94. 10.1016/j.jss.2014.08.062.

[8] Jun GT, Ward J, Clarkson PJ. Systems modelling approaches to the design of safe healthcare delivery: Ease of use and usefulness perceived by healthcare workers. Ergonomics 2010;53:829–47. 10.1080/00140139.2010.489653.

[9] Furniss D, Nelson D, Habli I, White S, Elliott M, Reynolds N, et al. Using FRAM to explore sources of performance variability in intravenous infusion administration in ICU: A non-normative approach to systems contradictions. Appl Ergon 2020;86. 10.1016/j.apergo.2020.103113.

[10] Chatzimichailidou MM, Ward J, Horberry T, Clarkson PJ. A Comparison of the Bow-Tie and STAMP Approaches to Reduce the Risk of Surgical Instrument Retention. Risk Analysis 2018;38:978–90. 10.1111/risa.12897.

[11] Andersen H, Røvik KA, Ingebrigtsen T. Lean thinking in hospitals: Is there a cure for the absence of evidence? A systematic review of reviews. BMJ Open 2014;4:1–8. 10.1136/bmjopen-2013-003873.

[12] Rah J-E, Manger R, Yock A, Kim G-Y. A comparison of two prospective risk analysis methods: Traditional FMEA and a modified healthcare FMEA. Med Phys 2016;43:3755–3755. 10.1118/1.4957534.

[13] Apkon M, Leonard J, Probst L, DeLizio L, Vitale R. Design of a safer approach to intravenous drug infusions: Failure mode effects analysis. Qual Saf Health Care 2004;13:265–71. 10.1136/qshc.2003.007443.

[14] Bradea I, Delcea C, Paun R. Healthcare Risk Management Analysis – A Bibliometric Approach. Journal of Eastern Europe Research in Business & Economics 2016;2015:1–11. 10.5171/2015.169472.

[15] Cure L, Zayas-Castro J, Fabri P. Challenges and opportunities in the analysis of risk in healthcare. IIE Trans Healthc Syst Eng 2014;4:88–104. 10.1080/19488300.2014.911786.

[16] Dückers M, Faber M, Cruijsberg J, Grol R, Schoonhoven L, Wensing M. Safety and risk management interventions in hospitals: A systematic review of the literature. vol. 66. 2009. 10.1177/1077558709345870.

[17] Walshe Kieran. Pseudoinnovation: The development and spread of. International Journal for Quality in Health Care 2009;21:153–9.

[18] Chiozza ML, Ponzetti C. FMEA: A model for reducing medical errors. Clinica Chimica Acta 2009;404:75–8. 10.1016/j.cca.2009.03.015.

[19] Nicolini D, Waring J, Mengis J. Policy and practice in the use of root cause analysis to investigate clinical adverse events: Mind the gap. Soc Sci Med 2011;73:217–25. 10.1016/j.socscimed.2011.05.010.

[20] Grant KP, Pennypacker JS. Project management maturity: An assessment of project management capabilities among and between selected industries. IEEE Trans Eng Manag 2006;53:59–68. 10.1109/TEM.2005.861802.

[21] Mullaly M. Longitudinal Analysis of Project Management Maturity. Project Management Journal 2006;36:26–73.

[22] Ward J, Clarkson J, Buckle P, Berman J, Lim R, Jun T. Prospective Hazard Analysis: Tailoring Prospective Methods to a Healthcare Context 2010:358. 10.1016/b978-0-12-398338-1.00011-7.

[23] Kubra G. Good risk assessment practice in hospitals 2018.

[24] Popp PL. Analysis of ASHRM membership survey on healthcare captives. J Healthc Risk Manag 2007;27:19–23. 10.1002/jhrm.5600270105.

[25] Kaya GK, Ward J, Clarkson J. A Review of Risk Matrices Used in Acute Hospitals in England. Risk Analysis 2019;39:1060–70. 10.1111/risa.13221.

[26] Kaya GK. A system safety approach to assessing risks in the sepsis treatment process. Appl Ergon 2021;94:103408. 10.1016/j.apergo.2021.103408.

[27] Kaya GK, Ward J, Clarkson J. Evaluation of risk management practices: data analysis of NHS England hospitals. Proceedings of the Global Joint Conference on Industrial Engineering and Its Application Areas 2016 2016.

[28] Simsekler MCE, Card AJ, Ward JR, Clarkson PJ. Trust-level risk identification guidance in the NHS East of England. International Journal of Risk and Safety in Medicine 2015;27:67–76 10.3233/JRS-150651.

[29] Simsekler MCE, Card AJ, Ruggeri K, Ward JR, Clarkson PJ. A comparison of the methods used to support risk identification for patient safety in one UK NHS foundation trust. Clin Risk 2015;21:37–46. 10.1177/1356262215580224.

[30] Bricknell M, Moore G. Health Risk Management Matrix - A Medical Planning Tool. J R Army Med Corps 2007;153:87–90.

[31] The British Standards Institution. BSI Standards Publication BS EN ISO 9000 : 2015 Quality management systems - Fundamentals and Vocabulary 2015:62.

[32] International Organization for Standardization. BS ISO 31000:2018 Risk management - guidelines. Switzerland: 2018.

[33] Casselman J, Onopa N, Khansa L. Wearable healthcare: Lessons from the past and a peek into the future. Telematics and Informatics 2017;34:1011–23. 10.1016/j.tele.2017.04.011.

[34] Federation of European Risk Management Associations. A Risk Management Standard. 2003.

[35] International Organization for Standardization. ISO/IEC 31010:2019 BSI Standards Publication Risk management – Risk assessment. Switzerland: 2019.

[36] British Standard Institute. (BS 31100) Code of Practice for Guidance for the Implementation of ISO: 31000 2011.

[37] ASHRM. Patient Safety Risk Managment Playbook. Chicago: ASHRM; 2015.

[38] VHA National Center for Patient Safety (NCPS). Guide to Performing a Root Cause Analysis 2020:52.

[39] VHA National Center for Patient Safety (NCPS). Healthcare Failure Modes and Affects Analysis (HFMEA) Guidebook 2021.

[40] Ball DJ, Watt J. Further Thoughts on the Utility of Risk Matrices. Risk Analysis 2013;33:2068–78. 10.1111/risa.12057.

[41] Card AJ, Ward J, Clarkson PJ. Successful risk assessment may not always lead to successful risk control: A systematic literature review of risk control after root cause analysis. J Healthc Risk Manag 2012;31:6–12. 10.1002/jhrm.20090.

